# PesTrapp mobile app: A trap setting application for real-time entomological field and laboratory study

**DOI:** 10.1101/2021.02.05.21251191

**Authors:** Cheong Yoon Ling, Rosilawati Rasli, Mohd Khairuddin Che Ibrahim, Siti Futri Farahininajua Fikri, Nur Ayuni Nazarudin, Lim Kuang Hock, Khairul Asuad Muhamed, Mohd Zahari Tajul Hassan, Mohd Izral Yahya Umpong, Mohd Zainuldin Taib, Nazni Wasi Ahmad, Lee Han Lim

**Affiliations:** Biomedical Museum Unit, Special Resource Centre, Institute for Medical Research, Ministry of Health Malaysia, Jalan Pahang, 50588 Kuala Lumpur, Malaysia; Medical Entomology Unit, Infectious Disease Research Centre, Institute for Medical Research, Ministry of Health Malaysia, Jalan Pahang, 50588 Kuala Lumpur, Malaysia; Biomedical Research, Strategic & Innovation Management Unit, Institute for Medical Research, Ministry of Health Malaysia, Level 6, Block C6, National Institutes of Health (NIH), Setia Alam Selangor, Malaysia

**Keywords:** Dengue, evaluation, ovitrap index, *Aedes aegypti*, *Aedes albopictus*

## Abstract

Zoonotic diseases such as malaria, dengue, Zika and chikungunya remain endemic in many countries. Setting and deploying trap to capture the host/vector species are fundamental to understand their density and distribution. Human effort to manage the trap data accurately and timely is an exhaustive endeavour when the study area expands and study period prolongs. One stop mobile app to manage and monitor the process of animal trapping, from field to laboratory level is still scarce. We developed a new mobile app named “PesTrapp” especially for medical entomologists to acquire the vector density index based on the mobile updates of ovitraps and species information in field and laboratory. This study aimed to highlight the mobile app’s development and design, describe the user experience and evaluate the preliminary user assessment of the mobile app. The mobile app was developed using mobile framework and database. User evaluation of the mobile app was based on the adjusted Mobile App Rating Scale and Standardized User Experience Percentile Rank Questionnaire. The process flows of system design and detailed screen layouts were described. The user experiences of using the app in a project to study *Aedes* surveillance in six study sites in Selangor, Malaysia were described. The overall mean user evaluation score of the mobile app was 4.0 (SD=0.6), which showed its acceptability by the users. The PesTrapp, a one-stop solution, is anticipated to improve the entomological surveillance work processes. This new mobile app can contribute as a tool in the vector control countermeasure strategies.

## INTRODUCTION

Emerging and re-emerging zoonotic diseases are significant issues in global health ^1^. Dengue, an important vector-borne zoonotic disease globally, is endemic in many tropical countries. In the continued absence of an effective vaccine and antivirals, chemical control, biological control, environmental management, health education and legislation are utilised to control the *Aedes* vectors and the diseases they carry ^2^. Ovitrap, a device which consists of a 300-ml plastic container with straight and slightly tapered sides ^3^ is used to characterise the temporal and spatial distribution and abundance of *Aedes* mosquitoes in a locality by taking advantage of the preference of *Aedes* mosquitoes laying eggs in containers. Usually, a minimal number of ovitraps, i.e. 50 to 100 ovitraps, deployed in one locality, indoor or/and outdoor, and recollected after 5 to 7 days can be used to estimate vector abundance in a large urban area (Mogi et al. 1990). The ovitrap index, the entomological indicator, which quantifies the infestation frequency of vector mosquitoes, is the number of traps with study subjects (mosquito species) divided by the total recovered traps. The mean number of larvae per trap is used to estimate the adult mosquito abundance. These data have been used to construct early warning prediction model to forestall dengue outbreaks so that remedial action can be taken to avoid and suppress the outbreak ^4,5^.

Currently, the ovitrap deploying process is tedious and resource hunger. The field team first deploys the ovitraps in a selected study site. While deploying in the study site, a distinct ovitrap code is assigned for each ovitrap and the house number, road and descriptions are recorded in a prescribed paper datasheet. To obtain the coordinates of the location, the same ovitrap code is inserted in the Global Positioning System (GPS) devices and the coordinates of the location are captured. After five to seven days, the team search and collect the traps based on the recorded information. The status of the ovitraps, whether missing or found, is updated in the datasheet. Back in the laboratory, the datasheet is then updated in a master Microsoft Excel file. The data from the GPS devices is exported to .csv or .dat file and updated in the master Excel file. Then, the species of the adult mosquitoes is identified and recorded in paper by the laboratory assistants if no computer is available in the laboratory. Subsequently, the species information will be matched and updated in the master Excel file. Finally, the ovitrap index and mean larva per trap are analysed and reported in graphs or charts. The common problems of these conventional manual activities are bulk data to compile, inconsistency of recording method, errors while transcribing the information and misidentification of ovitraps. Different platforms of data recording and storage, from paper, GPS devices to Excel sheet, are not integrated. When the data grows large with many site monitoring, without a database system, the data management and interpretation process with spreadsheets alone are exhaustive and error prone.

Several mobile applications are utilised to monitor animal trap by embedded sensor ^6,7^, analyse the images of pests on glue board ^8^ and monitor the capture-mark-recapture process ^9^. The US Centers for Disease Control and Prevention (CDC) developed a mobile app named “Epi Info Vector Surveillance Application” to track and enter mosquito surveillance data and insecticide resistance bioassay testing ^10^. The public commercialised outdoor GPS recording mobile app (e.g. Handy GPS, Geo Tracker, A-GPS Tracker) would be the alternatives to acquire coordinates of traps, but they are not integrated with the project database. As far as our knowledge can tell, mobile app to manage and monitor the process of animal trapping, from field to laboratory level, in particular for vector borne zoonotic disease studies is scarce. Based on the requests from the medical entomologists, a one stop project-based mobile application (app) that integrate all processes to obtain *Aedes* index, from field to laboratory, is urgently needed. Therefore, we developed a mobile app named “PesTrapp” as a trap setting application for real-time field and laboratory study. This study aimed at three objectives, firstly, to describe the development and design of the mobile app; secondly, to illustrate the user experience of using the mobile app; thirdly, to evaluate the preliminary user assessment of the mobile app.

## MATERIALS AND METHODS

### Development of the mobile app

The PesTrapp is developed from scratch using PHP ^17^ and MySQL database storage ^18^ on the Android operating system as Google’s Android is open source Linux based ^15^ and widely used ^16^. The user interface is developed using javascript code and html5. The final code are compiled and bundled using Apache Cordova ^19^, open source Onsen javacsript library ^20^ into wrapper and APK file. The services used by PesTrapp are Google Maps Application Programming Interface (API) for the mapping and Google Firebase API ^21^ for pushing notification.

### Design of the mobile app

The functions of PesTrapp mobile are sequentially divided into several processes, namely preparation, deployment of ovitraps, collection of ovitraps, identification of species and reports. Upper part of Fig. 1 showed the screen layout of preparation process. System administrator or project leader pre-register the PesTrapp users in the web dashboard. Subsequently, registered and login user able to register the study site in the mobile app by setting the site code, site name, type of site (landed houses or high-rise flats/apartments/condominiums), number of traps and code of traps. Lower part of Fig. 1 depicted the deployment of ovitraps process. There are four main functions in the main menu of the app, namely, deployment of ovitraps, collection of ovitraps, identification of species and reports. User clicks the “Deployment of ovitraps” button and search or select the site from list. For landed houses, user inserts trap status (indoor/outdoor), house number, road, description of traps, records coordinate and takes photo of trap location. For high-rise flats/apartments/condominiums, user inserts trap status (indoor/outdoor), house number, level, block, description of traps and takes photo. The information is instantly saved in the database server. Capturing of coordinates per ovitrap is impossible in high-rise as GPS signal is often low. The ovitrap status screen is displayed for both deployment and collection of ovitrap using color coding. Green color indicates ovitraps that have been deployed, white color indicates ovitraps that have not yet been deployed.

**Fig. 1.**
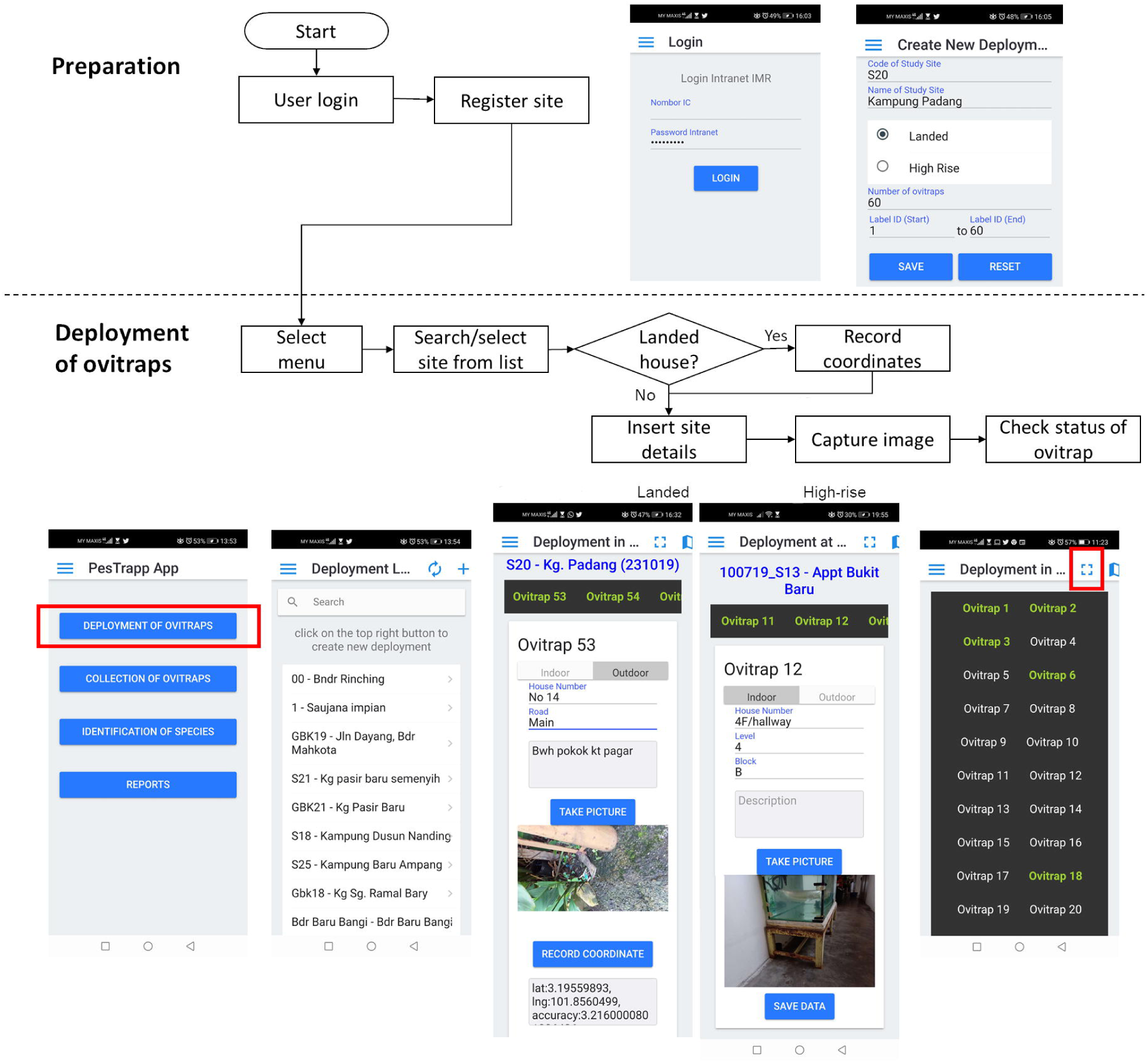
Screen layouts of the PesTrapp for preparation and deployment of ovitraps process.

Upon collecting the trap as shown in upper part of Fig. 2, user refers to the retrieved map and photo. User traces the location of ovitraps based on saved information and then updates status whether each ovitrap is recovered or not recovered. If the trap is missing or broken, user updates as “Not recovered” option. With the ovitrap status page, user knows the latest status of ovitrap being collected. Green colour indicates ovitraps that have been collected, white colour indicates ovitraps that have not yet been collected and red colour indicates not recovered trap. Lower part of Fig. 2 showed the identification of species process and screen layouts. In the laboratory, user inserts the species identification details directly in the PesTrapp. The app instantly displays overall, indoor and outdoor ovitrap index as well as overall, indoor and outdoor mean larvae per recovered ovitrap by species as shown in Fig. 3. User can export the data to .csv and print.

**Fig. 2.**
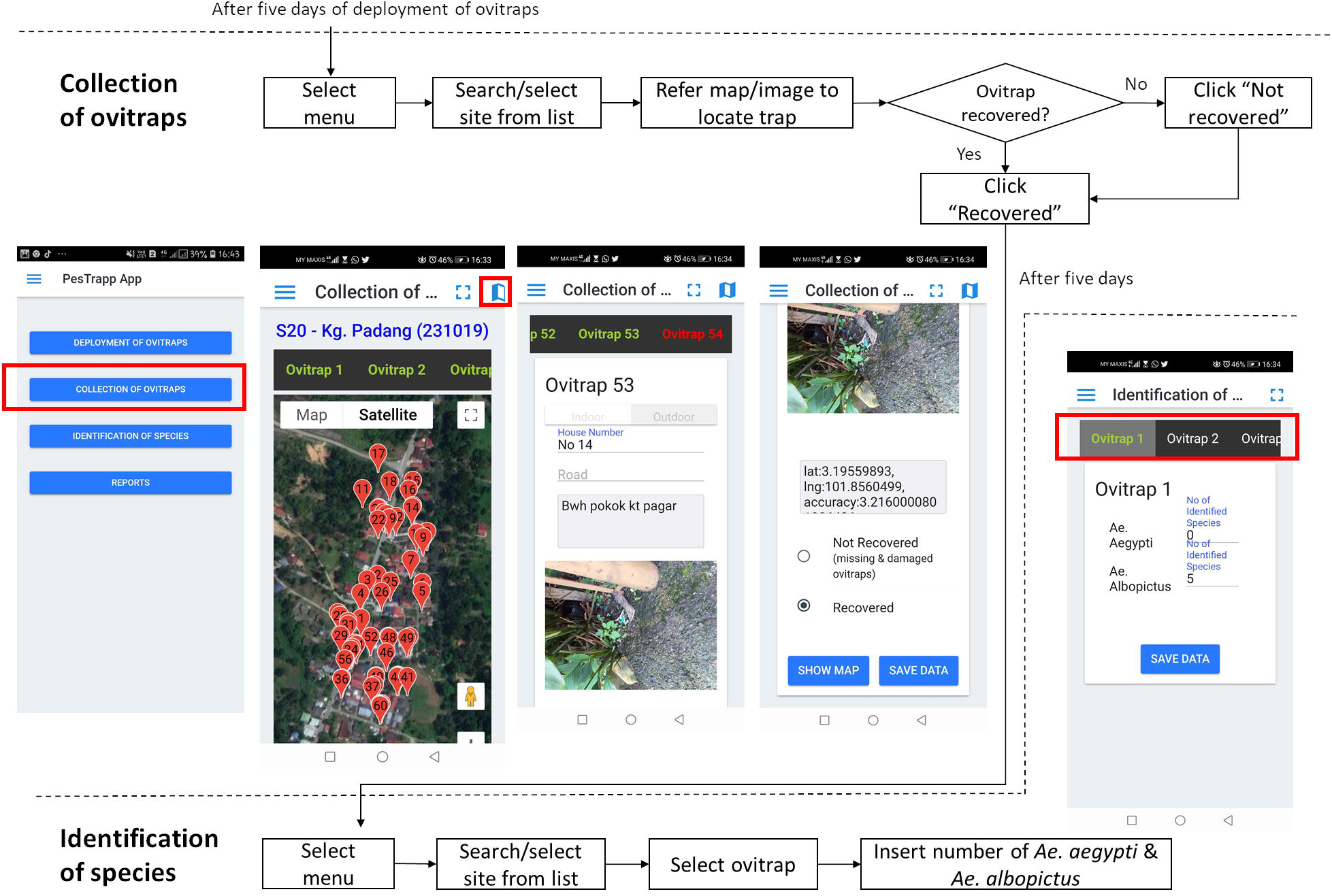
Screen layouts of the PesTrapp for collection of ovitraps and identification of species process.

**Fig. 3.**
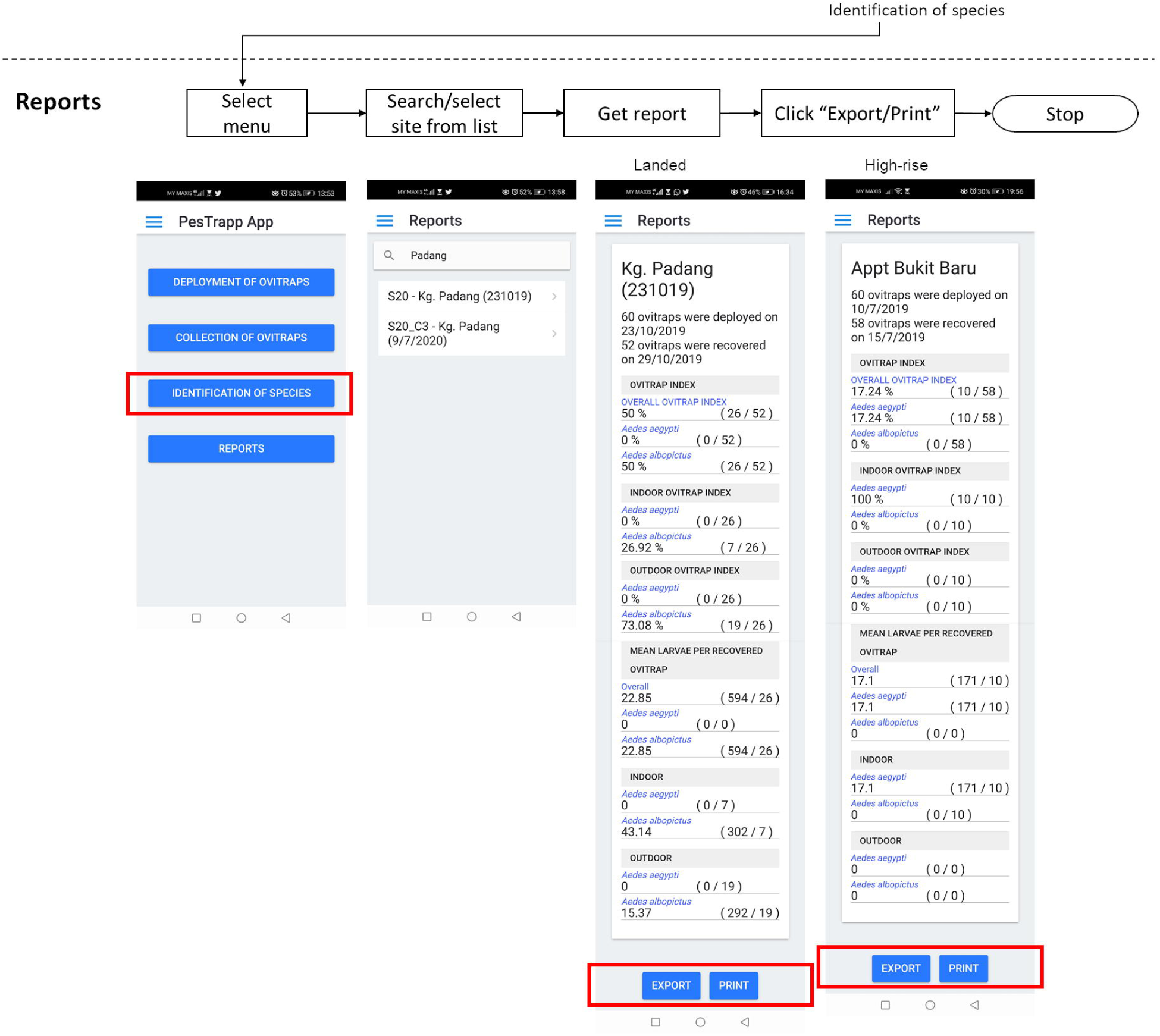
Screen layouts of the PesTrapp for reports process.

### Hands-on user field experiences

We described the field experience under real life situation of using the PesTrapp app based on the project Insecticide Resistance Mapping. Ovitraps deployment for *Aedes* sampling (60 ovitraps for each site) were carried out at six study sites in the district Gombak and Hulu Langat, Selangor. Five of the sites were landed houses (Kampung Sungai Chinchin, Kampung Baru Ampang, Kampung Padang, Kampung Jawa and Bandar Mahkota) and one was high-rise apartment (Apartment Bukit Baru).

### User evaluation of the app

We developed the evaluation form of 11 quantitative questions and three qualitative questions based on Mobile App Rating Scale (MARS) ^22^ and Standardized User Experience Percentile Rank Questionnaire (SUPR-Qm) ^23^. MARS multidimensionally assess the quality of mobile applications ^23^ and was applied to studies such as dengue reporting and mapping, health communication, and behavior modification mobile application ^24^ and gamified pain management mobile application ^25^. SUPR-Qm was initially created to capture user experience for website but was later adapted to measure user experience for mobile application ^23^. The evaluation form of 11 quantitative questions measured the engagement, functionality, aesthetic, information and acceptability of the PesTrapp mobile app users. Ten questions were adapted from MARS and one integrability question was extracted from SUPR-Qm. Each item adheres to 5-point scale (1-inadequate, 2-Poor, 3-Acceptable, 4-Good, 5-Excellent). Data were summarized as mean and standard deviation for each item and category that were then described accordingly. The evaluation form of three qualitative questions assessed the appreciation of app, usage of features and suggestion to improve the app.

## RESULTS

### User experience using the app

Project leader first added user login for the team members in the project. Site details and number of ovitraps deployed were entered accordingly in PesTrapp. It was found useful as early notification to medical entomologists’ team prior to placement of ovitrap in field. Home dwellers were well informed with the ovitrapping procedure. Two ovitraps - indoor and outdoor - were deployed at each house once permission was granted by the home dwellers. Following that, respective details of house address and description of located ovitraps were filled in accordingly. Once ovitraps were located outdoor and indoor, an image of deployed ovitrap was captured and location was recorded with the best accuracy level. All details were saved by clicking the “Save Data” button and then it is ready for next deployment of ovitraps. In total, 60 ovitraps were deployed per sites and were collected after 5 days in field for the oviposition process. Digital records of ovitrap location details and captured images during placement helped in tracking ovitraps during collection.

After five days of deployment in the field, the ovitraps were collected according to the recorded ovitrap data. By clicking through the “Collection of Ovitraps” button, the recorded study sites were listed in the collection listing. During the collection process, the location of the deployed ovitraps were confirmed through the captured images and ovitrap location coordinates on “Show Map” page. Recovered ovitraps were recorded as “Recovered”, while missing or tampered ovitraps were recorded as “Not Recovered”. The number of recovered ovitraps varied according to sites, for i.e. Gombak district, 58/60 ovitraps and 54/60 ovitraps were collected at Apartment Bukit Baru and Kampung Sungai Chinchin respectively. Meanwhile for Hulu Langat district, 57/60 ovitraps, 52/60 ovitraps, 56/60 ovitraps and 50/60 ovitraps were collected at Kampung Baru Ampang, Kampung Padang, Kampung Jawa and Bandar Mahkota respectively. The collected ovitraps were then brought back to the laboratory for identification of species process.

Collected mosquito larvae of L3 stage were identified using compound microscope after five days of collection. Identification listing based on study sites were automatically generated in the PesTrapp by clicking the “Identification of Species” button. The identified larvae were pooled according to their species of either *Aedes aegypti* or *Aedes albopictus* and the number of collected larvae were recorded in the PesTrapp. The total number of recovered ovitraps, ovitrap index and mean larvae per recovered ovitrap for both indoor and outdoor by species were automatically generated in the “Report” section in the PesTrapp. Overall, ovitrapping method that involves collection and placement of ovitraps in field; and processing ovitraps in laboratory were conducted in a timely manner using PesTrapp mobile application.

### Preliminary user evaluation of the app

Seven project users who used the PesTrapp mobile app evaluated the mobile app on 21 July 2020 after using the app for at least one month. Table 1 showed the profiles of users as 7 young adults (four female, three male) from age 24 to 36, with a mean age of 27.3 years (*SD=4*.*1 years*) and one of them was the project principal investigator. Most users used project phone to run the app one or more times per week (71%) and worked in both field and laboratory work (86%). The months of experience in project were mixed with the mean of 24.3 months (*SD=16*.*2*).

**Table 1.** Sample Characteristics.

| Variable | Frequency | Percent |
| --- | --- | --- |
| Gender |  |  |
| Female | 4 | 57% |
| Male | 3 | 43% |
| Project role |  |  |
| Principal investigator | 1 | 14% |
| Co-investigator | 0 | 0% |
| Research assistant | 6 | 86% |
| Use app on |  |  |
| Own phone | 2 | 29% |
| Project phone | 5 | 71% |
| Work scope |  |  |
| Field work | 1 | 14% |
| Laboratory work | 0 | 0% |
| Both field and laboratory work | 6 | 86% |
|  | Mean | SD |
| Age | 27.3 | 4.1 |
| Months in project | 24.3 | 16.2 |

The mean evaluation score was 4.0 (*SD=0*.*6*) and ranged from 3.4 to 4.6 out of 5.0. The 2-item “information quality” subscale assesses the achievable goals and visual explanation of concepts through images and maps, scored the highest with a mean of 4.4 (*SD=0*.*6*) as shown in Table 2. The 2-item “acceptability” subscale scored a mean of 4.2 (*SD=0*.*8*). All users would recommend this app to people who might benefit from it. Most of the users (4/7) rated the app four stars and above, while some users (3/7) rated the app three stars. Next, the “aesthetic” subscale assesses the layout of mobile app obtained the mean of 3.9 (*SD=0*.*7*). Concerning the 2-item “engagement” subscale with the mean of 3.8 (*SD=1*), target group scored way higher than the interactivity, with the mean of 4.1 (*SD=0*.*9*) and mean of 3.4 (*SD=1*.*1*) respectively. Regarding the 4-item “functionality” subscale, with similar score to engagement, users evaluated higher for ease of use, with the mean of 4.1 (*SD=0*.*9*) than the performance of with the mean of 3.7 (SD=1), navigation with the mean of 3.6 (SD=1.3) and integrability with the mean of 3.7 (SD=0.8).

**Table 2.** Evaluation of item scores for PesTrapp application (n=7).

| Subscale | Score Mean (SD) |
| --- | --- |
| **Engagement** | **3.8 (1)** |
| Target group | 4.1 (0.9) |
| Interactivity | 3.4 (1.1) |
| **Functionality** | **3.8 (1)** |
| Ease of use | 4.1 (0.9) |
| Performance | 3.7 (1) |
| Navigation | 3.6 (1.3) |
| Integrability | 3.7 (0.8) |
| **Aesthetic** | **3.9 (0.7)** |
| Layout | 3.9 (0.7) |
| **Information quality** | **4.4 (0.6)** |
| Goals | 4.3 (0.8) |
| Visual Information | 4.4 (0.5) |
| **Acceptability** | **4.2 (0.8)** |
| Recommendation | 4.6 (0.5) |
| Rating | 3.8 (0.9) |

The qualitative evaluation of users on the appreciation of the app were positive. Most users praised the app for its efficiency in ovitrap monitoring especially the features of image capturing and coordinates. *“Effective and time saving. Very useful to trace back the ovitraps based on photos taken in field and recorded coordinates”. “Effectiveness and well help in tracking the ovitrap”. “Ease the data collecting process and reduce the usage of paper”. “Easy to use when collecting ovitraps during outdoor. Ease the process to find by looking at picture taken*.*”*

All users reported frequently usage of the features, namely “deployment of ovitrap” and “collection of ovitraps” in the app. *“The deployment and collection of ovitraps as I am involved in a lot on fieldwork”*. One of the users also frequently used the “identification of species” feature. *“Recording the location ovitrap in field and picture. Record larvae species”*.

Users suggested few aspects to improve the app. Majority of users suggested the “edit” button (5 of 7) and “back” button (4 of 7) to be added in the app. *“Add button edit and delete to make sure the list of localities is move effective and easily to trace”. “Edit button”. “Add back button”*. Two of the seven users requested the photo upload features for reporting purpose. *“(1) option to upload photos from photo gallery (2) Edit button on homepage so can make amendment at the key-in data (3) ‘back’ button on every page, instead of keep going back to the home page”*. The other two users preferred the “delete” button to be added too. *“Add up edit, delete, back button and also add up block option for the high-rise building”*. One of the users suggested the need for smooth interface for file management.

## Discussion

A mobile app to monitor the deployment of ovitraps, collection of ovitraps, identification of species and reporting is developed. The mobile app development process required the full commitment from the medical entomologists, system analysts and mobile developers. The main drive to the development of a comprehensive mobile app depends on to what extent the system analysts and mobile developers understand the problem of current practices and to what extent the users or stakeholders explain or highlight the problem. This is in line with Pandey et al. who commented the process of mobile app models should be explanatory, as much as possible ^26^. Continuous discussion and review are essential. From our experience, system analysts and mobile developers constantly had meetings and dialogues with the medical entomologists to understand existing limitations of current practices, conceive, design, develop and evaluate the prototypes. The prototype is then used and tested by the project members during field and laboratory works.

The advancement in the internetworking and touch screen phone maximize the capability of mobile app technology ^27^. The centralised database manages bulk data in split seconds. Monitoring of ovitraps with paper and not integrated are not recommended when the data grows big with repetitive monitoring and large coverage extent. PesTrapp is specially designed to integrate the mobile app technology with centralised database server via internet. The data integrity is ensured even if the works in field or laboratory are handled by different personnel. The data lost issue at individual desktop is avoided by storing data centrally in dedicated and isolated database server away from existing project site due to improper handling of data, on-site fire disaster and virus-infected desktop. The mobile apps with centralised database are widely adopted by other mobile app studies such as mosquito vector surveillance ^28^, sentinel sites reporting ^29^ and disease symptoms surveillance ^30^.

The entomological field and laboratory work progress are easily monitored by project manager with the mobile app. The different colour representations in the ‘ovitrap status’ page serve as work progress updates for the project manager. The completed (green), uncompleted (white) and not recovered (red) representations inform the real-time status of the trap whether it has been deployed or collected. The data transparency and forensic are guaranteed with the information that provided by the timestamps and user location when a task is added, modified, completed; when the app is installed and last launched in the device ^31^. In this case, users are unable to tamper and modify the date of deployment, collection and species identification.

The user evaluation obtained a high overall score and in most of its subscales. The mobile app is well-accepted by the users that they would recommend this mobile app to people who might benefit from it. All users agreed that the visual explanation of concepts with images and maps were mostly clear, logical and correct. Most users found the app easy to use and the content information were well-targeted and appropriate. The functions, buttons and menus of the app worked timely. The functionality and engagement of the app were generally accepted by the users. However, on “navigation”, three users took some time and effort to move between screens uninterrupted. This is in line with the qualitative question 3 in which majority users suggested to add “back” button in the app for easy navigation. The “interactivity” subscale scored the lowest due to the availability of only basic interactive features of user input and notification. Most users suggested to add “edit” button in the app in the qualitative question 3. Hence, mobile developer added the functions based on the fed-back from users.

The features of the PesTrapp improve user experience through visual graphic presentation. Deploying trap is easy in the field, but collecting trap requires good recall memory after five to seven days of deployment. The image and maps serve as a better graphical presentation for the user to recall the location of trap, in line with the studies that discovered geographical map increases recall of text ^32,33^ and graphic enhances perceptual and cognitive processes ^34^ supported this idea. Besides, the real-time notification of alert ‘saved’ messages confirms the interaction and responsiveness of the mobile app.

Protection of the security, privacy and confidentiality of the mobile app users should be ensured. The users should be educated on the formal mobile policy including authentication, integrity and data transfer ^35^. The server should be regularly updated with the latest security control. The mobile developers should minimize the mobile data access permissions to avoid malicious virus such as spyware, Rootkit and Trojan horses ^36^. The users should keep mobile operating system up to date, encrypt mobile phone and use mobile security software ^37^. To protect the privacy of the residents, image of the position of trap should not be captured with the residents and inside the house without permission from the residents. The access to the data should be protected with password.

The future directions to improve the app are the customization possibilities to other animal trap setting studies and integration with other technologies. PesTrapp is potential to be customised for different animal trapping studies such as flies, rats and squirrels by adjusting the number of animal species and density index. Further research to integrate PesTrapp with other technologies, ranging from the digital eggs counting ^38^, larval image identification ^39^, species identification ^40^, QR code/barcode ^41^ and climate data ^42,43^ are anticipated to improve and extend its capabilities. Nevertheless, there should be a balance of cost and benefit in applying new technologies to the existing low-cost intervention.

PesTrapp is a potential tool to gather species density data nationwide. Usually, in the entomological studies, mosquito density index is segregated for local study area for evaluating vector control interventions and larval survey during outbreak ^44–46^. Gathering the mosquito density index gives a better understanding of the distribution and movement of mosquitoes for macro- and micro-habitat studies. This mobile app is potential to be the data gathering and managing tool towards big data era, likewise the social media gather user data for business intelligence ^47^ and web questionnaire gather research data for subject matter analysis ^48,49^. If PesTrapp is to be applied nationwide, its database could be linked to the disease surveillance system and open source climate repository for integrating multiple ends - entomology, epidemiology and climate for dengue transmission prediction ^50^ for better control measures to manage disease hotspot and handle outbreak.

There are some limitations of this mobile app. Firstly, PesTrapp is developed for Android version phone, but not iOS phone. We are targeting Android phone as it is preferred by the users with a larger market share of around 85.4% in year 2020 ^16^. Secondly, the GPS satellites signal is undetectable inside building and generally poor in high-rise building. We designed the capture of coordinates differently for the landed and high-rise building. For high-rise, coordinates of ovitraps were not mandatory due to poor signal of GPS, whereas for landed houses, each ovitrap should be reported its coordinates. Nevertheless, record of levels and blocks are essential for high-rise building.

## Conclusions

PesTrapp exhibits the potential to transform conventional trapping method to real-time trap managing intervention. The commitment from both users and developers in translating problem-solving ideas into solution, designing, implementing and testing the proposed app is essential. With the real-time trap setting and collecting mobile app, centralised and organized managing of the density of species contribute to strategic site selection for future vector control interventions. Furthermore, the potential integration of the mobile app with other digital platforms in vector control including disease surveillance, fogging monitoring system, disease hotspot mapping system will enable better vector control management in handling outbreak. Future works will be the enhancement of the features of mobile app and customization of the application to other potential animal trap studies.

## Data Availability

The datasets used and analysed during the current study are available from the corresponding author on reasonable request.

## List of abbreviations

GPS: Global Positioning System
CDC: Centers for Disease Control and Prevention
app: application
ITEX: International Invention, Innovation & Technology Exhibition, Malaysia
FR: functional requirements
NFR: non-functional requirements
UI: user interface
API: Application Programming Interface
MARS: Mobile App Rating Scale
SUPR-Qm: Standardized User Experience Percentile Rank Questionnaire

## Acknowledgments

We would like to thank the Director General of Health Malaysia and the director, Institute for Medical Research (IMR) for their permission to publish this article. This work was supported by Ministry of Health Research Grant (NMRR-18-12-39653). Special appreciation is extended to all Medical Entomology Unit staff for their support during this study.

## Authors’ contributions

CYL RR MKCI designed the study together. CYL and RR drafted the manuscript and finalised it with MKCI SFFF NAN MZT NWA LHL provided feedback on the manuscript. MKCI developed the mobile application with the assistance from MZTH. RR led the field and laboratory works and worked with SFFF NAN KAM MIYU in data collection, data assurance and coordination of the study. MKCI CYL led the mobile app testing and evaluation. LKH and CYL analysed and interpreted the user questionnaire data. All authors have read and approved the final manuscript.

## Ethics approval and consent to participate

Ethical approval protocol from the Medical Research and Ethics Committee (MREC), Ministry of Health Malaysia was followed (reference number KKM/NIHSEC/P18-668(4)). Written informed consent was obtained from users who consented to take part in the survey.

## Consent for publication

Not applicable.

## References

1. Kilpatrick AM, Randolph SE. Drivers, dynamics, and control of emerging vector-borne zoonotic diseases. Lancet. 2012;380(9857):1946–1955. doi:https://doi.org/10.1016/S0140-6736(12)61151-9

2. Chang MS, Christophel EM, Gopinath D, et al. Challenges and future perspective for dengue vector control in the Western Pacific Region. West Pacific Surveill response J WPSAR. 2011;2(2):9–16. doi:10.5365/WPSAR.2010.1.1.012

3. Lee HL. Aedes ovitrap and larval survey in several suburban communities in Selangor Malaysia. Mosquito-Borne Dis Bull. 1999;9:9–15.

4. Mogi M, Choochote W, Khamboonruang C, Suwanpanit P. Applicability of presence–absence and sequential sampling for ovitrap surveillance of Aedes (Diptera: Culicidae) in Chiang Mai, northern Thailand. J Med Entomol. 1990;27(4):509–514.

5. Lee HL. Sequential sampling: its application in ovitrap surveillence f Aedes (Diptera: Culicidae) in Selangor, Malaysia. Trop Biomed. 1992;9:29.

6. Eiran B-D, Earle DM. Small animal trap. February 2017.

7. Loughlin D. Developments in the world of insect detection. Int Pest Control. 2013;55(2):88.

8. Freudenberg JR, Diller CD, Smyth KS. Analyzing images of pests using a mobile device application. September 2019.

9. Bateman HL, Lindquist TE, Whitehouse R, Gonzalez MM. Mobile application for wildlife capture–mark–recapture data collection and query. Wildl Soc Bull. 2013;37(4):838–845.

10. Global Health D of PD and M. Epi Info Vector Surveillance Application. Centers for Disease Control and Prevention. https://www.cdc.gov/parasites/education_training/epi_info_app.html. Published 2017. Accessed October 7, 2020.

11. Gupta V, Chopra RK, Chauhan DS. Status of Non-Functional Requirements in Mobile Application Development: An Empirical Study. J Inf Technol Res. 2017;10(1):59–84.

12. Peischl B, Ferk M, Holzinger A. The fine art of user-centered software development. Softw Qual J. 2015;23(3):509–536. doi:10.1007/s11219-014-9239-1

13. Olsina L, Becker P. Linking business and information need goals with functional and non-functional requirements. In: Proceed. of the XXI Conferencia Iberoamericana En Software Engineering (CIbSE’18), Bogotá, Colombia, Published by Curran Associates.; 2018:381–394.

14. Chung L, Nixon BA, Yu E, Mylopoulos J. Non-Functional Requirements in Software Engineering. Vol 5. Springer Science & Business Media; 2012.

15. Alliance OH. Android. https://www.openhandsetalliance.com/android_overview.html. Published 2010. Accessed May 5, 2020.

16. Chau, Melissa; Reith R. Smartphone Market Share. IDC Corporate USA. https://www.idc.com/promo/smartphone-market-share/os. Published 2020. Accessed July 7, 2020.

17. Group TP. PHP. https://www.php.net/. Published 2001. Accessed April 3, 2020.

18. Corporation O. MySQL. https://www.mysql.com/. Accessed July 8, 2020.

19. Foundation AS. Apache Cordova. https://cordova.apache.org/. Published 2020. Accessed May 4, 2020.

20. Team M x OU. Onsen UI. https://onsen.io. Accessed July 8, 2020.

21. Developers G. Google Firebase Cloud Messaging. https://firebase.google.com/products/cloud-messaging. Published 2020. Accessed July 5, 2020.

22. Stoyanov SR, Hides L, Kavanagh DJ, Zelenko O, Tjondronegoro D, Mani M. Mobile App Rating Scale: A New Tool for Assessing the Quality of Health Mobile Apps. JMIR mHealth uHealth. 2015;3(1):e27. doi:10.2196/mhealth.3422

23. Sauro J, Zarolia P. SUPR-Qm: a questionnaire to measure the mobile app user experience. J Usability Stud. 2017;13(1):17–37.

24. Karita T, Francisco ME, Watanabe K. An Integrated mHealth App for Dengue Reporting and Mapping, Health Communication, and Behavior Modification: Development and Assessment of Mozzify. JMIR Form Res. 2020;4(1):e16424.

25. Hoffmann A, Faust-Christmann CA, Zolynski G, Bleser G. Toward Gamified Pain Management Apps: Mobile Application Rating Scale–Based Quality Assessment of Pain-Mentor’s First Prototype Through an Expert Study. JMIR Form Res. 2020;4(5):e13170.

26. Pandey M, Litoriya R, Pandey P. Validation of existing software effort estimation techniques in context with mobile software applications. Wirel Pers Commun. 2020;110(4):1659–1677.

27. Tracy K. Mobile Application Development Experiences on Apple’s iOS and Android OS. Potentials, IEEE. 2012;31:30–34. doi:10.1109/MPOT.2011.2182571

28. Lozano-Fuentes S, Ghosh S, Bieman JM, et al. Using Cell Phones for Mosquito Vector Surveillance and Control. In: SEKE.; 2012:763–767.

29. Lwin MO, Jayasundar K, Sheldenkar A, et al. Lessons from the implementation of Mo-Buzz, a mobile pandemic surveillance system for dengue. JMIR public Heal Surveill. 2017;3(4):e65.

30. Stanton MC, Mkwanda SZ, Debrah AY, et al. Developing a community-led SMS reporting tool for the rapid assessment of lymphatic filariasis morbidity burden: case studies from Malawi and Ghana. BMC Infect Dis. 2015;15(1):214. doi:10.1186/s12879-015-0946-4

31. Azfar A, Choo K-KR, Liu L. Forensic taxonomy of android productivity apps. Multimed Tools Appl. 2017;76(3):3313–3341.

32. Kulhavy RW, Stock WA, Kealy WA. How geographic maps increase recall of instructional text. Educ Technol Res Dev. 1993;41(4):47–62.

33. Yen J, Lee C, Chen I. The effects of image-based concept mapping on the learning outcomes and cognitive processes of mobile learners. Br J Educ Technol. 2012;43(2):307–320.

34. Winn W. Contributions of perceptual and cognitive processes to the comprehension of graphics. In: Advances in Psychology. Vol 108. Elsevier; 1994:3–27.

35. Martínez-Pérez B, De La Torre-Díez I, López-Coronado M. Privacy and security in mobile health apps: a review and recommendations. J Med Syst. 2015;39(1):181.

36. Zhu H, Xiong H, Ge Y, Chen E. Mobile app recommendations with security and privacy awareness. In: Proceedings of the 20th ACM SIGKDD International Conference on Knowledge Discovery and Data Mining.; 2014:951–960.

37. Ahvanooey MT, Li Q, Rabbani M, Rajput AR. A survey on smartphones security: software vulnerabilities, malware, and attacks. arXiv Prepr arXiv200109406. 2020.

38. Brasil LM, Gomes MMF, Miosso CJ, da Silva MM, Amvame-Nze GD. Web platform using digital image processing and geographic information system tools: a Brazilian case study on dengue. Biomed Eng Online. 2015;14(1):69. doi:10.1186/s12938-015-0052-2

39. Muñoz JP, Boger R, Dexter S, Low R. Mosquitoes and Public Health: Improving Data Validation of Citizen Science Contributions Using Computer Vision. In: Delivering Superior Health and Wellness Management with IoT and Analytics. Springer; 2020:469–493.

40. Sharma N, Colucci-Gray L, Siddharthan A, Comont R, van der Wal R. Designing online species identification tools for biological recording: the impact on data quality and citizen science learning. Roberts D, ed. PeerJ. 2019;6:e5965. doi:10.7717/peerj.5965

41. Boob A, Shinde A, Rathod D, Gaikwad A. Qr Code based mobile app and business process Integration. Int J Multidiscip Curr Res. 2014;2:1014–1017.

42. Cheong YL, Burkart K, Leitão PJ, Lakes T. Assessing weather effects on dengue disease in Malaysia. Int J Environ Res Public Health. 2013;10(12). doi:10.3390/ijerph10126319

43. Jain R, Sontisirikit S, Iamsirithaworn S, Prendinger H. Prediction of dengue outbreaks based on disease surveillance, meteorological and socio-economic data. BMC Infect Dis. 2019;19(1):272. doi:10.1186/s12879-019-3874-x

44. Ali R, Azmi RA, Ahmad NW, et al. Entomological Surveillance Associated with Human Zika Cases in Miri Sarawak, Malaysia. Am J Trop Med Hyg. 2020;102(5):964–970.

45. Rosilawati R, Lee HL, Nazni WA, et al. Pyrethroid resistance status of Aedes (Stegomyia) aegypti (Linneaus) from dengue endemic areas in Peninsular Malaysia. IIUM Med J Malaysia. 2017;16(2).

46. Rozilawati H, Tanaselvi K, Nazni WA, et al. Surveillance of Aedes albopictus Skuse breeding preference in selected dengue outbreak localities, peninsular Malaysia. Trop Biomed. 2015;32(1):49–64.

47. Goncalves M, Cornelius Smith E. Social media as a data gathering tool for international business qualitative research: opportunities and challenges. J Transnatl Manag. 2018;23(2-3):66–97. doi:10.1080/15475778.2018.1475181

48. van Gelder MMHJ, Bretveld RW, Roeleveld N. Web-based Questionnaires: The Future in Epidemiology? Am J Epidemiol. 2010;172(11):1292–1298. doi:10.1093/aje/kwq291

49. Gosling SD, Vazire S, Srivastava S, John OP. Should we trust web-based studies? A comparative analysis of six preconceptions about internet questionnaires. Am Psychol. 2004;59(2):93.

50. Manogaran G, Lopez D. Disease surveillance system for big climate data processing and dengue transmission. In: Climate Change and Environmental Concerns: Breakthroughs in Research and Practice. IGI Global; 2018:427–446.

